# Systematic review of natural language processing (NLP) applications in magnetic resonance imaging (MRI)

**DOI:** 10.1101/2024.07.21.24310760

**Authors:** Gazal Mahmeed, Dana Brin, Eli Konen, Girish N Nadkarni, Eyal Klang

**Affiliations:** Division of Diagnostic Imaging, Sheba Medical Center, Ramat Gan, Israel; Faculty of Medicine, Tel Aviv University, Tel Aviv, Israel; Division of Data-Driven and Digital Medicine, Department of Medicine, Icahn School of Medicine at Mount Sinai, New York, NY, USA; The Charles Bronfman Institute of Personalized Medicine, Icahn School of Medicine at Mount Sinai, New York, NY, USA

## Abstract

**Background:** As MRI use grows in medical diagnostics, applying NLP techniques could improve management of related text data. This review aims to explore how NLP can augment radiological evaluations in MRI.

**Methods:** We conducted a PubMed search for studies that applied NLP in the clinical analysis of MRI, including publications up to January 4, 2024. The quality and potential bias of the included studies were assessed using the QUADAS-2 tool.

**Results:** Twenty-six studies published between April 2010 and January 2024, covering more than 160k MRI reports were analyzed. Most of these studies demonstrated low to no risk of bias of the NLP. Neurology was the most frequently studied specialty, with twelve studies, followed by musculoskeletal (MSK) and body imaging. Applications of NLP included staging, quantification, and disease diagnosis. Notably, NLP showed high precision in tumor staging classification and structuring of free-text reports.

**Conclusion:** NLP shows promise in enhancing the utility of MRI. However, there is a need for prospective studies to further validate NLP algorithms in real-time clinical and operational scenarios and across various radiology specialties, which could lead to broader applications in healthcare.

## Introduction

Natural language processing (NLP) combines computer science, artificial intelligence, and linguistics to improve how computers and humans interact. The introduction of technologies like ChatGPT in 2022 marked a major shift in NLP, showcasing its broad potential^1^.

In radiology, traditionally reliant on computer vision^2, 3^ NLP introduces a new angle^4, 5^, with many potential uses, including flagging findings, prioritizing patients, generating imaging protocols, and conducting research^6, 7^.

MRI, known for its high contrast resolution and no radiation, is becoming more prevalent in diagnostic practices^8^. Using NLP in MRI interpretation could enhance workflows, diagnostics, and patient care.

This review assesses how NLP improves MRI applications by enhancing textual analysis in radiology.

## Methods

This systematic review was reported according to the preferred reporting items for systematic reviews guidelines (PRISMA). The study is registered under PROSPERO, number (CRD42024518710).

### Search strategy

We searched literature to find studies on NLP’s clinical uses in MRI. The search was conducted on January 4, 2024, using the PubMed database.

Search keywords included “MRI”, “Magnetic resonance imaging”, “MRE”, “Magnetic resonance enterography”, “ NLP”, “Natural Language Processing“, “LLM”, “large language models”, and “chatGPT”. Details on complete search strategies are provided in *(**Supplementary Material**)*.

Inclusion criteria were studies that (1) evaluated the clinical applications of NLP for MRI, (2) original articles in english (3) articles exclusively pertaining to MRI imaging.

We excluded (1) non - available full-text articles, (2) written in language other than english, (3) studies that included various imaging modalities other than MRI, (4) not focusing on NLP techniques, (5) studies focused on image processing and interpretation rather than text-based data analysis.

### Study selection

Two reviewers (GM, DB) independently screened the titles and abstracts to determine whether the studies met the inclusion criteria. The full-text article was reviewed when the title met the inclusion criteria or when there was any uncertainty. Disagreements were adjudicated by a third reviewer (EK).

### Data extraction

We used a standardized sheet to collect data on publication year, study design, location, database size, criteria, NLP methods, radiology field, MRI technique, NLP usage in MRI context, and performance.

### Quality assessment and risk of bias

Quality was assessed by the adapted version of the Quality Assessment of Diagnostic Accuracy Studies (QUADAS-2) criteria ^9^. Details on quality assessment are provided in *(**Supplementary Material**)*.

### Data synthesis and analysis

The analysis in this review is mainly qualitative. The heterogeneity of the studies in the literature evaluating NLP in MRI, their methods and the reported results precludes us from performing a meta-analysis.

## Results

### Study selection and characteristics

The initial search yielded 823 articles, with 26 meeting our inclusion criteria. **Figure 1** summarizes the characteristics of the included studies. The studies were published between 2010 and 2024. **Table 1** lists the publications reporting on the use of NLP in MRI.

**Figure 1:**
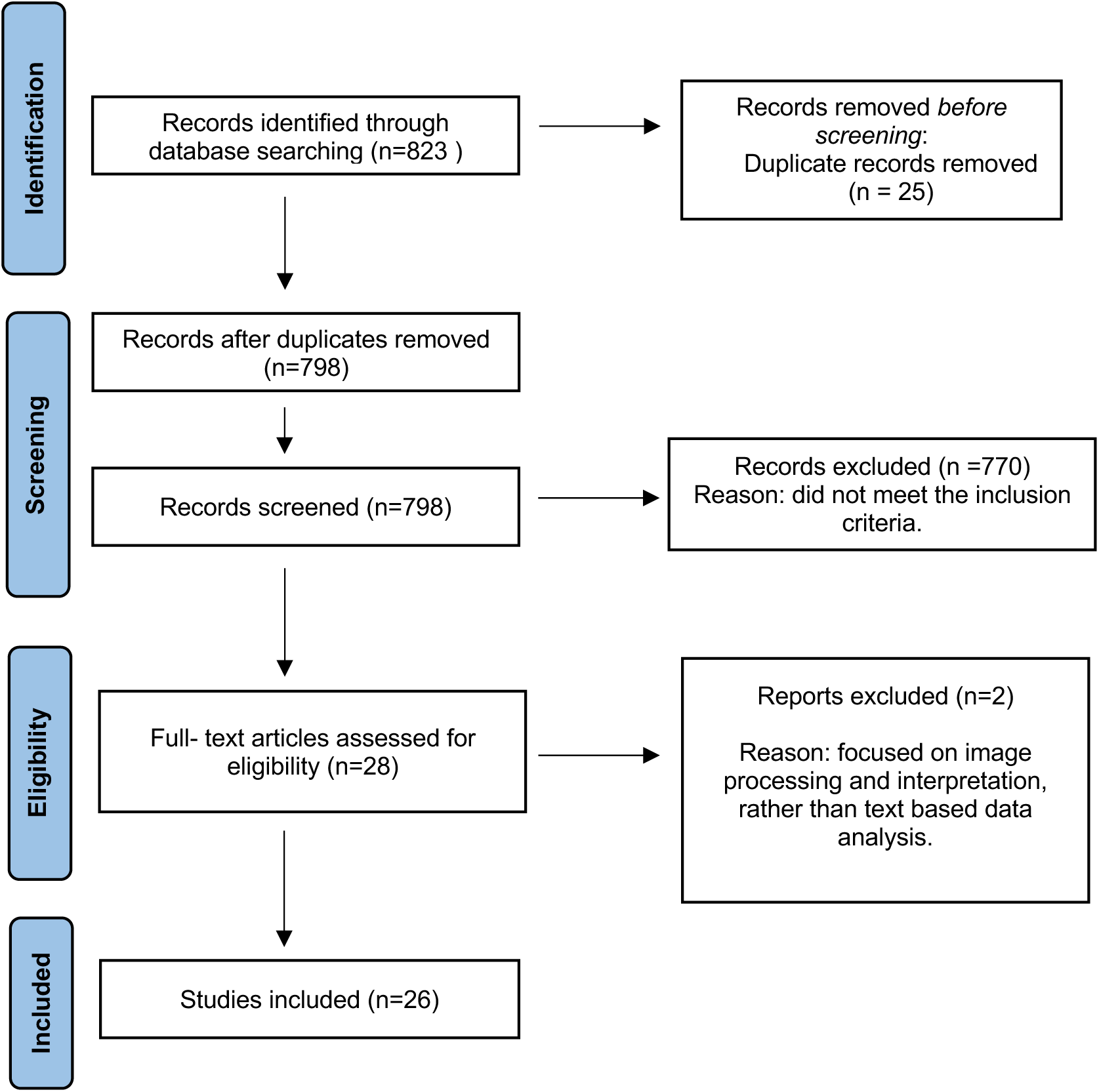
PRISMA flowchart of the study selection process.

**Table 1:**
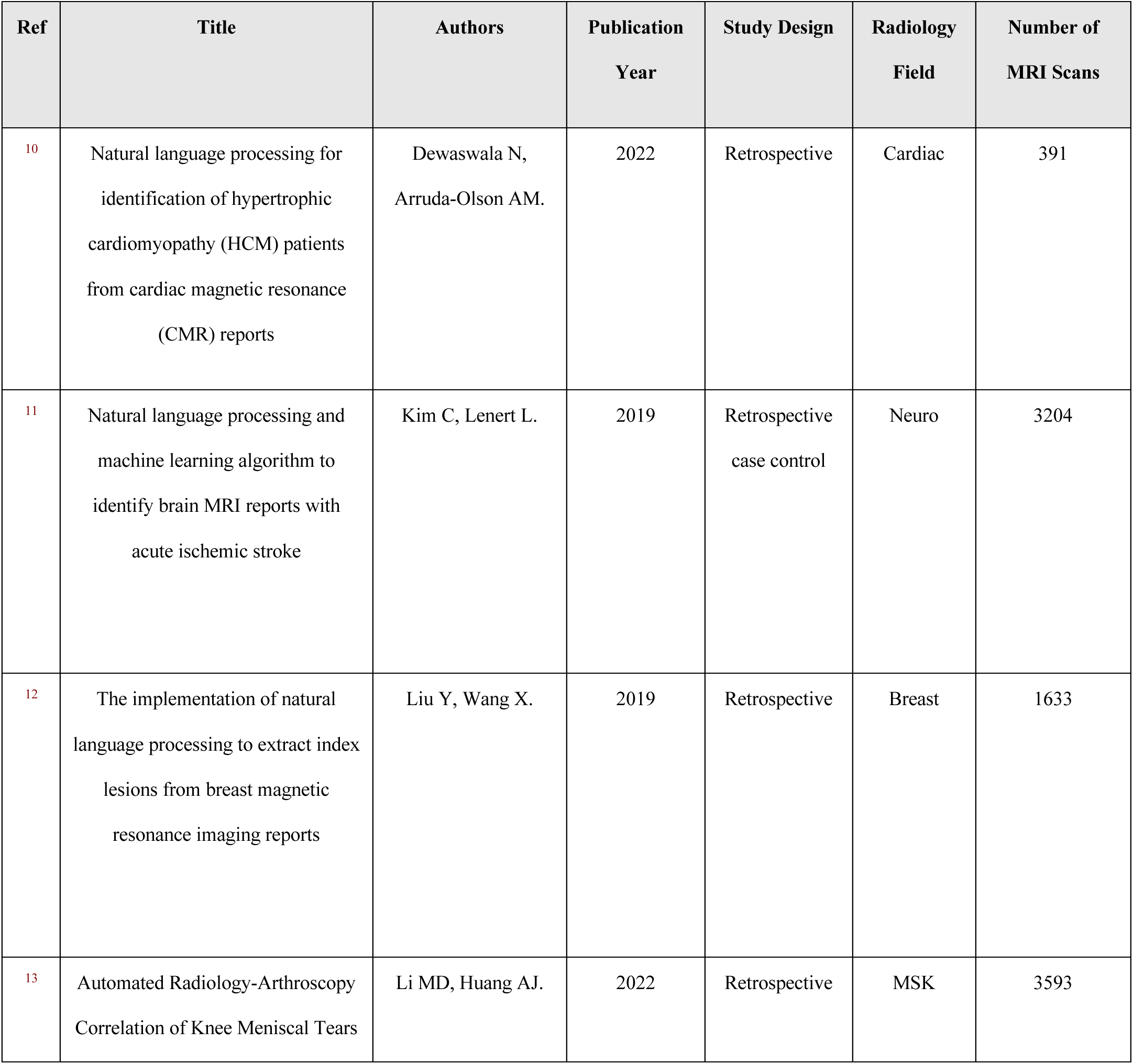
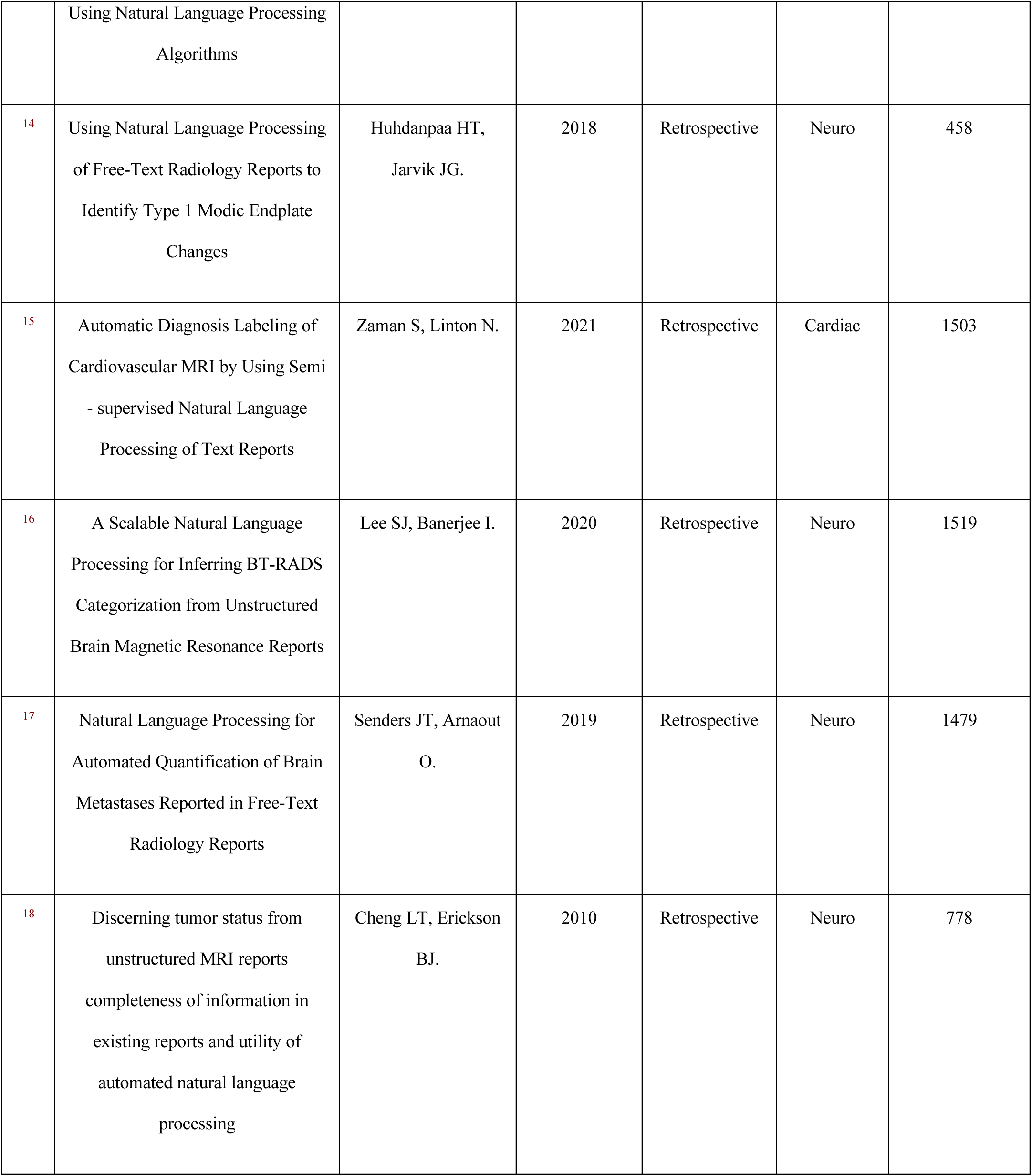
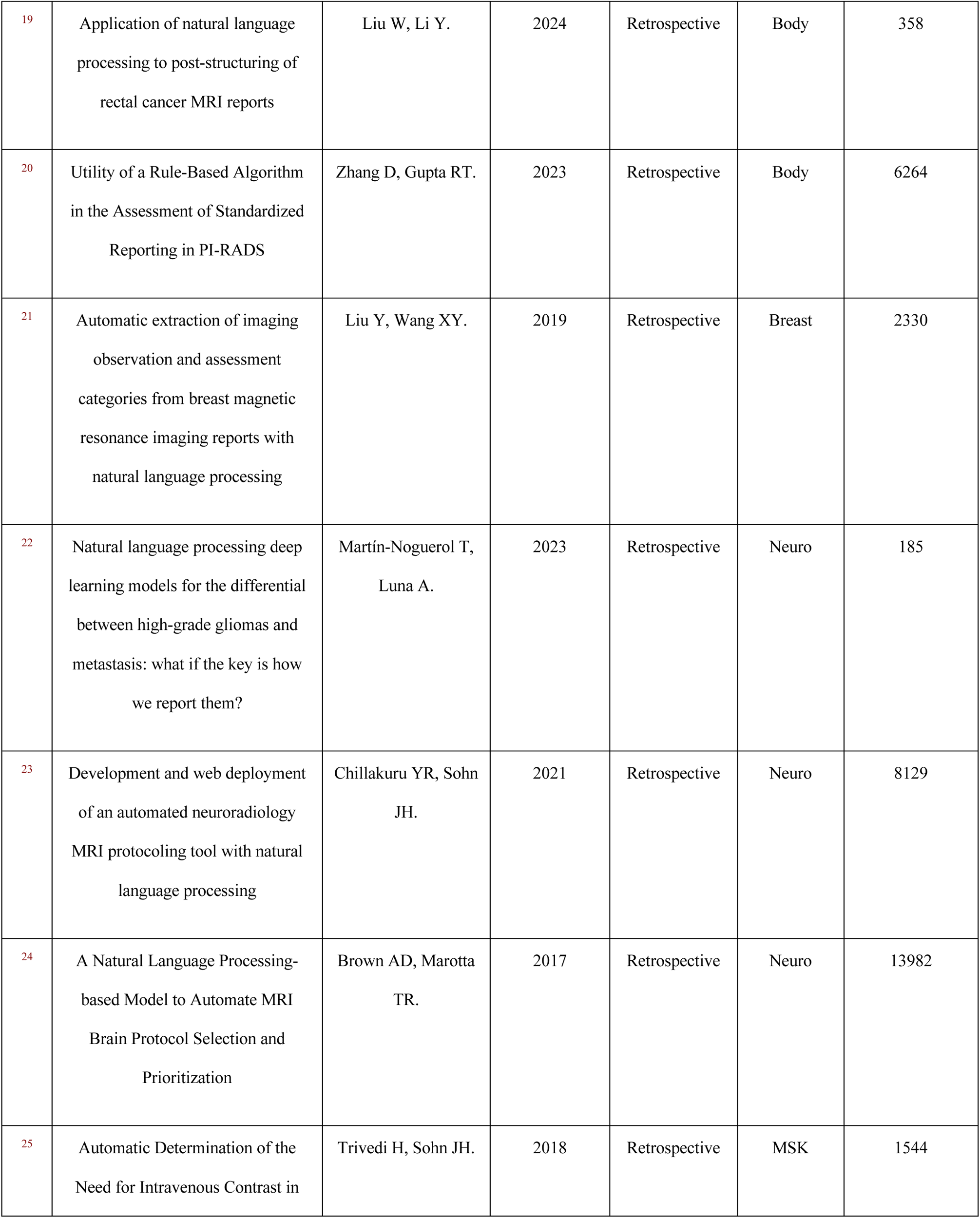
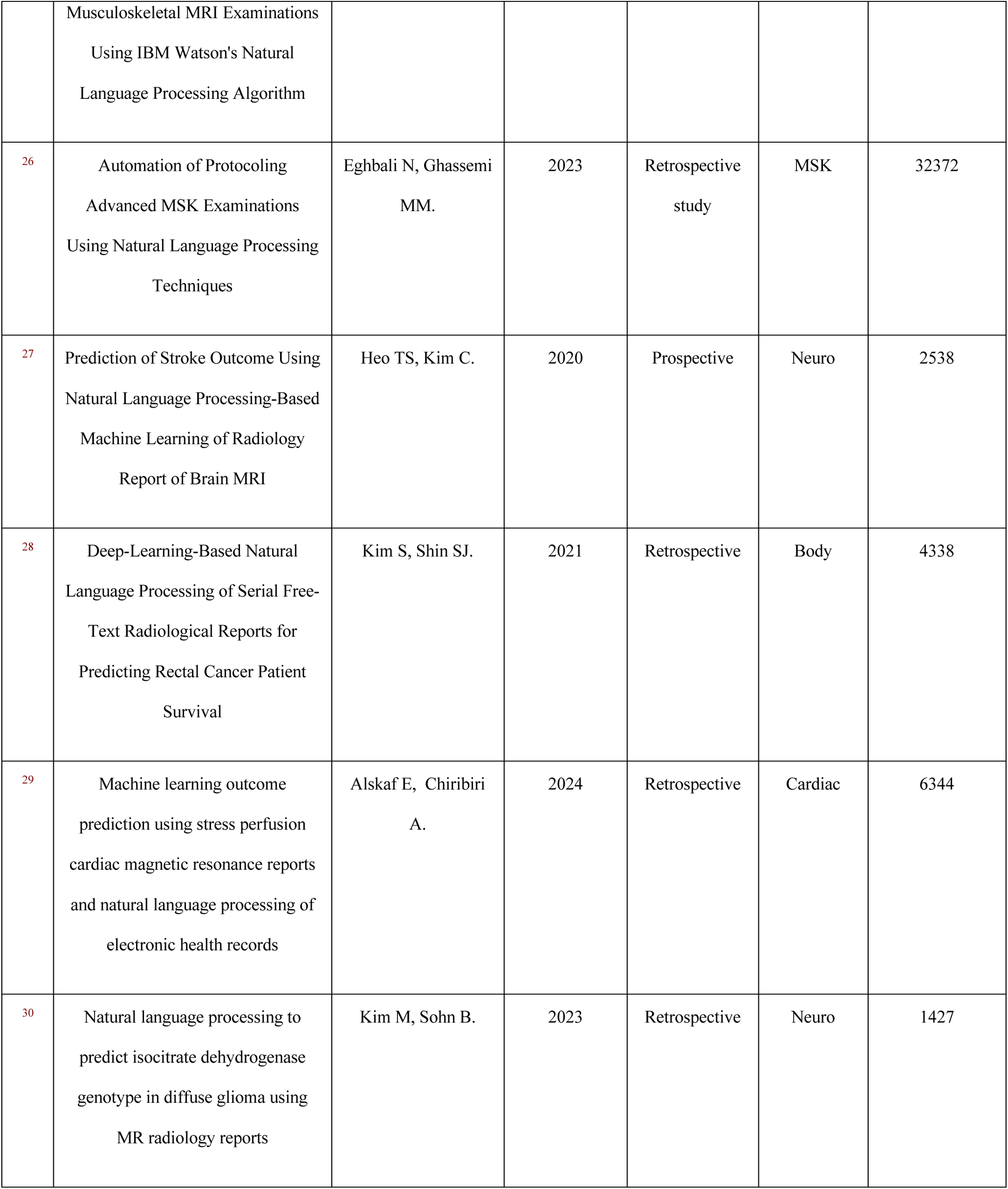
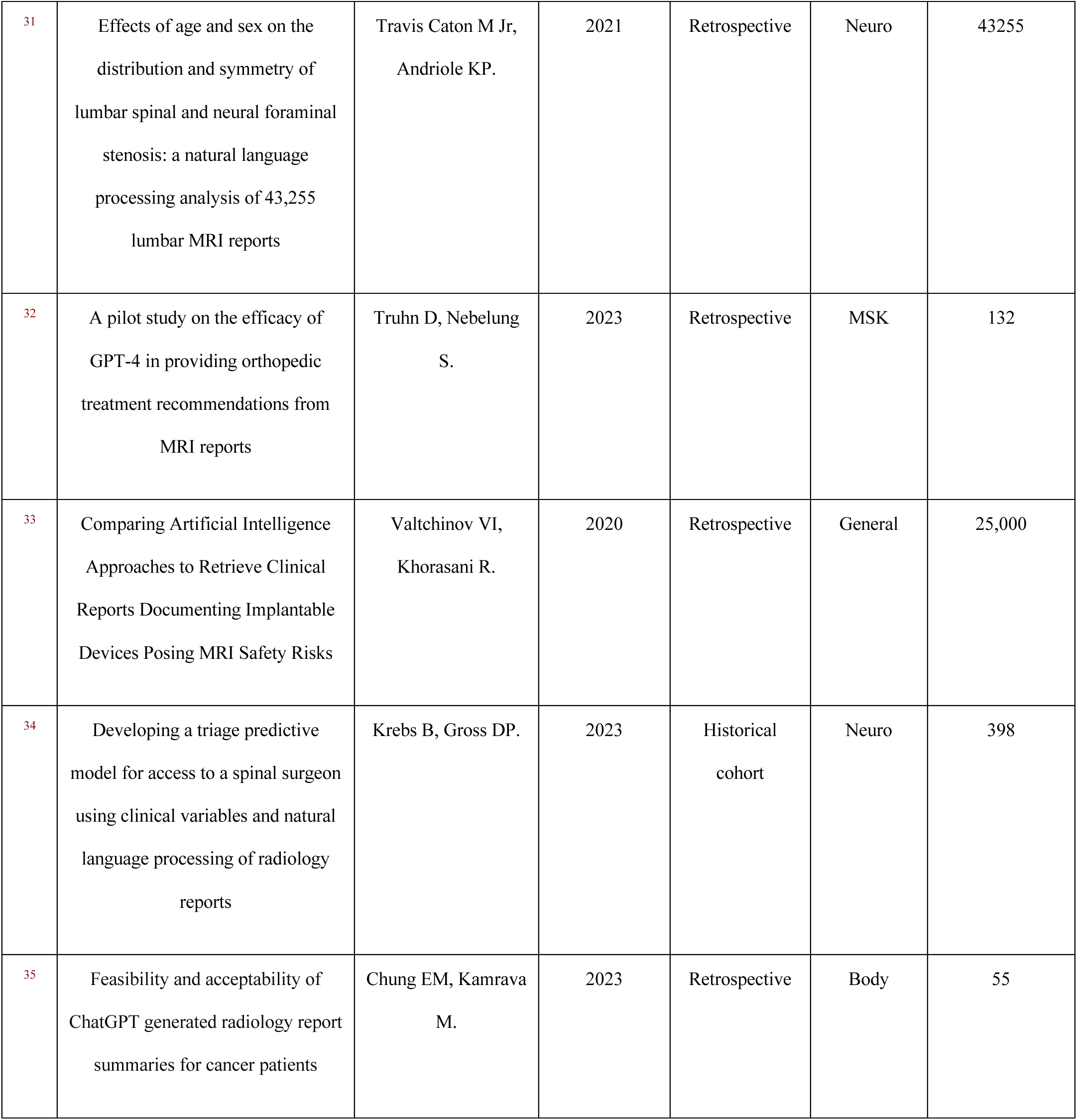
Publications reporting on the use of NLP in MRI.

### Descriptive summary of results

Over the past years there has been an increase in the number of publications of NLP applied to MRI.

**Figure 2** represents this general trend, with a peak of publications in 2023. All studies included were retrospective in nature, encompassing 163,209 MRI reports.

**Figure 2:**
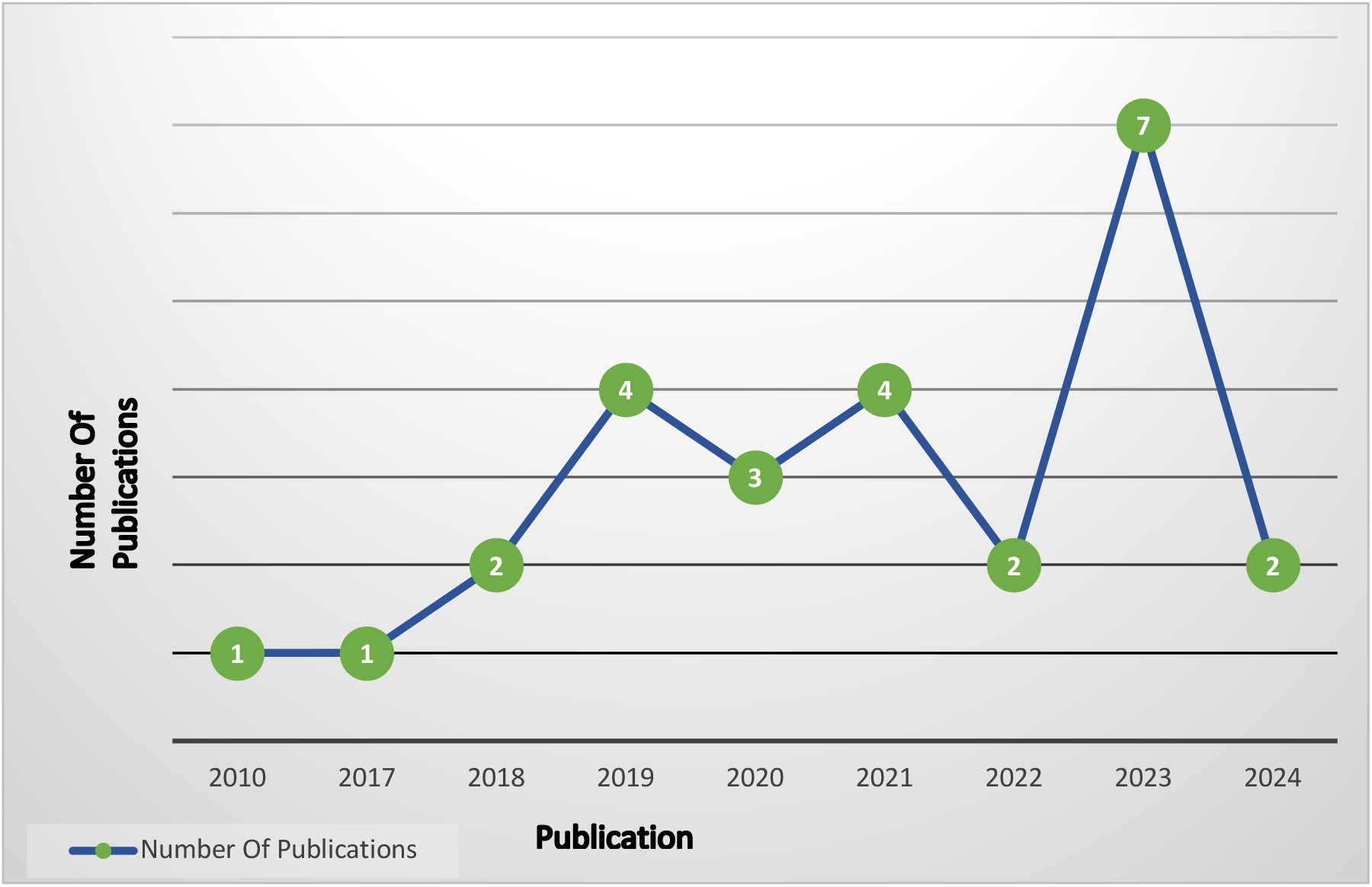
Evaluation of NLP applications in MRI research: trend analysis over time. Note that the studies included were published before January 2024.

**Figure 3** presents the distribution of radiology fields in the study. Neurology was the most prominent field, accounting for 12 out of 26 of the reviewed studies ^11^ ^14^ ^16^ ^17^ ^18^ ^22^ ^23^ ^24^ ^27^ ^30^ ^31^ ^34^. Musculoskeletal ^13 25^ ^26^ ^32^ and body ^19^ ^20^ ^28^ ^35^ imaging were studied in four publications each, making them the second most represented fields. Cardiac imaging followed with three research studies ^10^ ^15^ ^29^, while two studies involved breast imaging ^12^ ^21^. Additionally, there was one study that discussed the use of NLP for MRI in a more general context, without focusing on a specific field ^33^.

**Figure 3:**
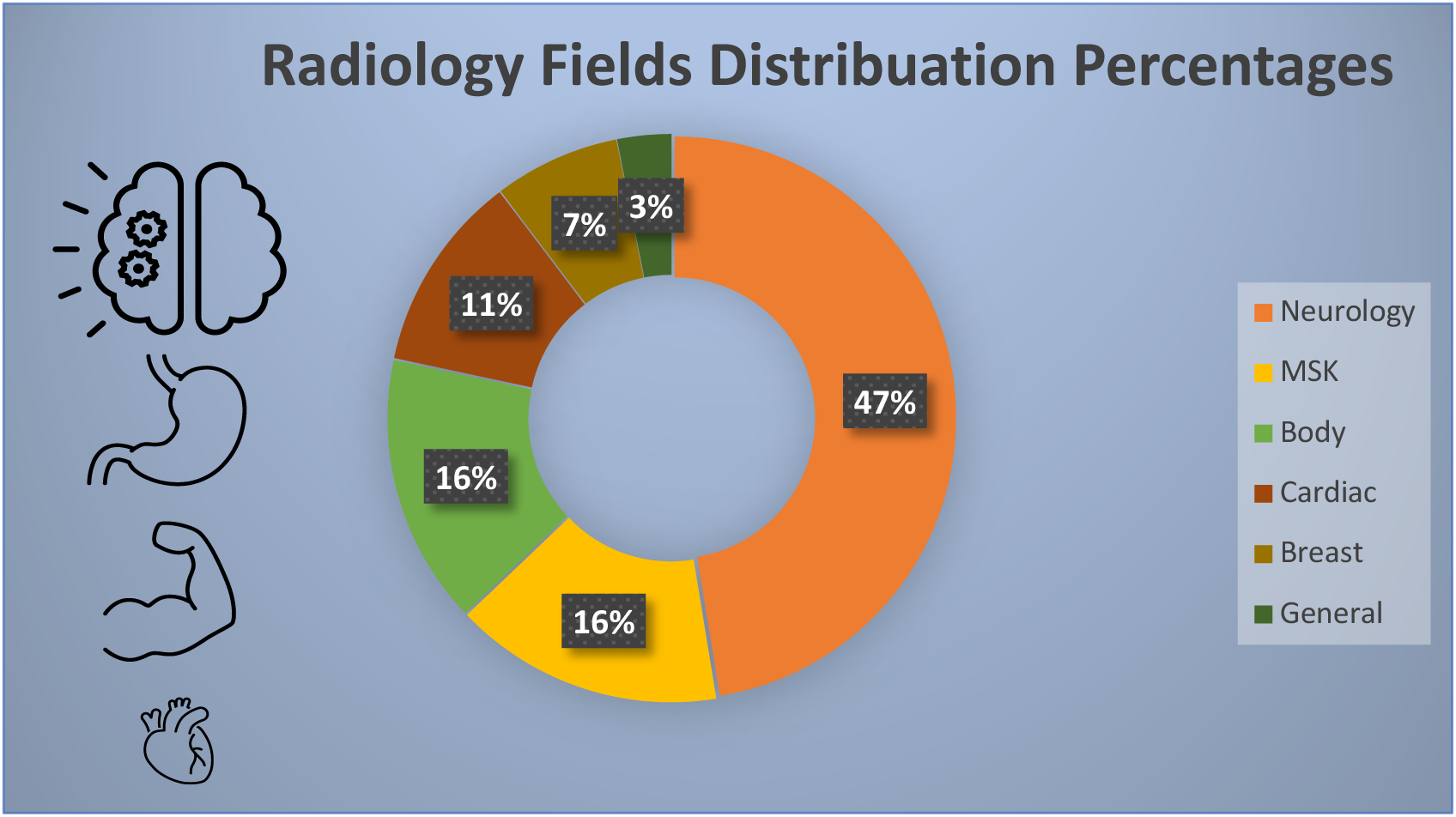
Radiology Fields Distribution Percentages.

The studies included various NLP techniques, such as rule based approaches, machine learning, and deep learning, including large language models (LLMs). The complete list of NLP techniques is detailed in **Table 2**.

**Table 2:** Summary of Studies Applying NLP to MRI: Modalities, Methods, Models, Tasks, and Performance Scores.

| Ref | NLP Clinical Application | MRI Modality | NLP Technique | NLP Task | Performance Score |
| --- | --- | --- | --- | --- | --- |
| <sup>10</sup> | Disease diagnosis | Cardiac | Rule based and machine learning | Data structuring | Accuracy 99% |
| <sup>11</sup> | Disease diagnosis | Brain | Comparison between multiple NLP models | Data structuring | Single decision tree showed the highest performance:<br>F1-measure 93.2%<br>Accuracy 98% |
| <sup>12</sup> | Disease diagnosis | Breast | Rule-based method | Data structuring | Identification of index lesion:<br>Recall and Precision > 85.0% |
| <sup>13</sup> | Disease diagnosis | Knee | Supervised machine learning models (logistic regression, SVM, and random forest) | Data structuring | Medial meniscus F1 scores 93%-94%<br>Lateral meniscus F1 scores 86%- 88% |
| <sup>14</sup> | Disease diagnosis | Spine | Rule-based NLP algorithm | Data structuring | Recall 70%<br>Specificity 99%<br>F1 79% |
| 15 | Disease diagnosis | Cardiac | Machine learning models | Data structuring | The BERT-based model achieved a micro-averaged F1 score 86% |
| 16 | Staging and quantification | Brain | Deep learning | Data structuring | Unstructured reports-f1 score 72%<br>structured reports-f1 score of 98% |
| 17 | Staging and quantification | Brain | Comparison between multiple NLP models | Data structuring | LASSO regression model demonstrated the best overall performance<br>AUC of 92%<br>Accuracy 83%<br>libration Intercept 6% |
| 18 | Staging and quantification | Brain | Statistical and rule-based methods | Data structuring | Sensitivity 80.6%<br>Specificity 91.6% |
| 19 | Staging and quantification | Rectum | Rule-based NLP model | Data structuring | Pre-2015 reports:<br>accuracy 93.8%<br>precision 95.6%<br>recall 87.1%<br>F1 score 91.2% |
|  |  |  |  |  | Post-2021 reports<br>accuracy 92.5%<br>precision 98.5%<br>, recall 94.15%<br>F1 score 96.3% |
| 20 | Staging and quantification | Prostate | Rule-based NLP model | Data structuring | Accuracy 92.6%<br>Precision 88.8%<br>Recall 85.6%<br>F1 score 87% |
| 21 | Staging and quantification | Breast | An internally developed NLP program | Data structuring | Recall 78.5%<br>Precision 86.1% |
| 22 | Staging and quantification | Brain | Deep learning models<br>CNN, BiLSTM,<br>BERT | Data structuring | CNN network provided<br>the best results :<br>Macro-avg precision<br>87.3%<br>Sensitivity 87.5%<br>F1 score 87.2% |
| 23 | Protocol selection | Spine, Brain | machine learning models (FastText, XGBoost) | Data structuring | Spine MRI model:<br>Accuracy 83.4%<br>AUC 88%. |
|  |  |  |  |  | The head MRI :<br>Accuracy 85.4%<br>AUC 94%<br>contrast brain protocol<br>AUC 92% |
| 24 | Protocol selection | Brain | Machine learning models (random forest, support vector machine (SVM), and k-nearest neighbor (KNN).) | MRI protocols | Accuracy:<br>Protocol selection 82.9%<br>Contrast administration<br>83.0%<br>Prioritization tasks 88.2% |
| 25 | Protocol selection | MSK, Spine | Comparison between multiple NLP models | MRI protocols | Watson vs. original protocol Sensitivity 74%<br>Accuracy 83%<br>Watson vs. second reader Sensitivity 81%<br>Accuracy 88%<br>Watson vs. original and second reader agreed case only reader Sensitivity 83%<br>Accuracy 90% |
| 26 | Protocol selection | MSK | Comparison between multiple NLP models | MRI protocols | Accuracy 83%<br>AUC 87% |
| 27 | Prognosis prediction | Brain | Machine learning models | Prediction models | RF algorithm had the best<br>AUC 78% |
| 28 | Prognosis prediction | Rectum | Deep learning | Prediction models | N/A |
| 29 | Prognosis prediction | Stress perfusion cardiac magnetic resonance | Machine learning models | Prediction models | support vector machine (SVM) was the best:<br>F1 score 24%<br>AUC 80% |
| 30 | Pathology prediction | Brain | Comparison between multiple deep learning models (LSTM , BiLSTM, BERT , BERT GCN , and BioBERT) | Prediction model | BERT GCN showed the highest performance:<br>AUC 85%-95%<br>CI 81% -89% |
| 31 | Comparative analysis | Lumbar spine | Rule-based natural language processing | Data structuring | Random sample of 100 LMRI reports<br>Accuracy 94.8%<br>At individual levels,<br>Accuracy ranged from 86.% |
|  |  |  |  |  | at right L5-S1 to 100% in<br>5/18 level instances<br>27.8%. |
| 32 | Clinical decision<br>support | Knee, Shoulder | Large language<br>models (chatGPT) | Treatment<br>recommendations | N/A |
| 33 | Safety protocol<br>compliance | Multiple | Expert-driven NLP;<br>ontology-driven NLP. | MRI protocols | MRI-Red:<br>- Ontology-Derived:<br>- Sensitivity: 96%<br>- Specificity: 90%<br>- Accuracy: 91%<br>MRI-Yellow:<br>- Ontology-Derived:<br>- Sensitivity: 76%<br>- Specificity: 62%<br>- Accuracy: 66% |
| 34 | Treatment<br>recommendations | Spine | N/A | Prediction models | Nagelkerke R – squared<br>R2 = 20% |
| 35 | MRI report summary | Prostate | Large language<br>models (chatGPT) | Data structuring | N/A |

We identified numerous valuable clinical applications of NLP in MRI. These applications are summarized in Figure 4, The key findings from each study are extensively described in **Table 3**.

**Figure 4:**
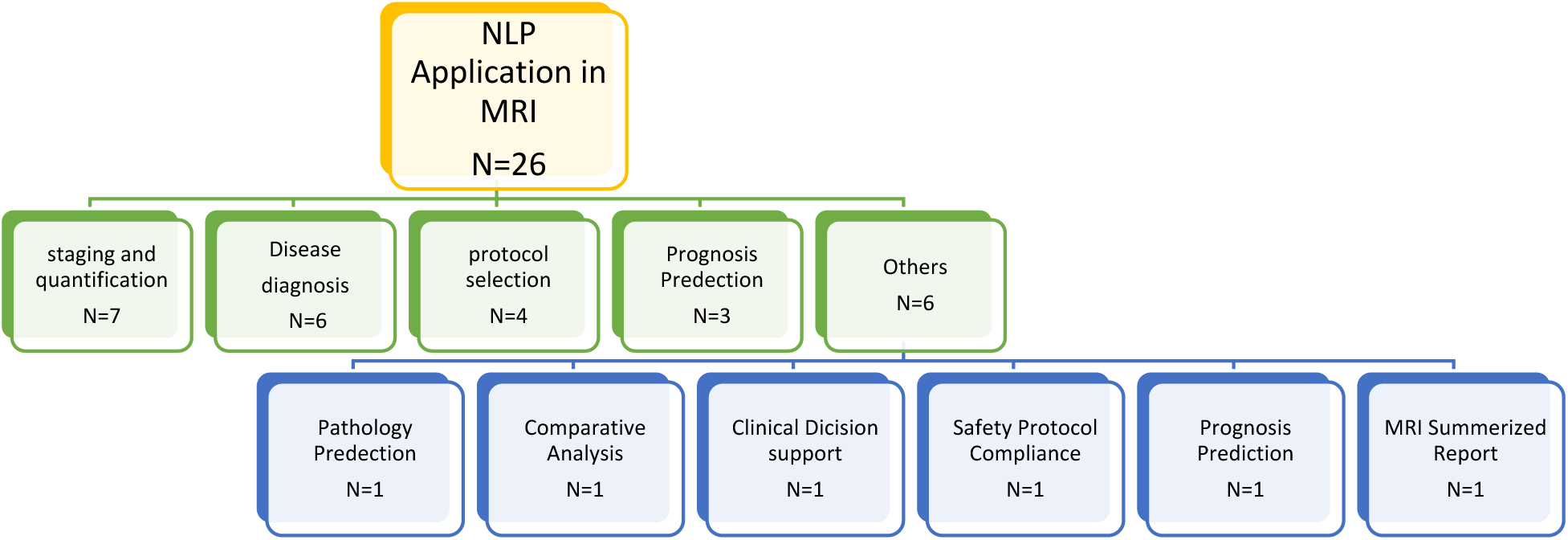
Natural language processing (NLP) MRI applications.

**Table 3:** Summary of Main Results and Limitations of Synthesized Studies.

| Ref | Main Findings | Study Limitations |
| --- | --- | --- |
| 10 | NLP identified and classified HCM from CMR narrative text reports with high performance. | 1. Complex sentences, ambiguity, and Sentences recorded in incorrect sections of the report were reasons for false-positive results.<br>2. The NLP algorithms used were developed and tested in a single tertiary medical center in a cohort of patients with suspected HCM. |
| 11 | Supervised ML based NLP algorithms are useful for automatic classification of brain MRI reports for identification of acute ischemic stroke (AIS) patients. Single decision tree was the best classifier to identify brain MRI reports with AIS. | 1. The text corpus was created at a single institution.<br>2. Only included brain MRI reports with conventional stroke MRI sequence. In clinical practice, full conventional brain MRI sequence could vary depending on the degree of emergency in each situation, the patient's condition, and the laboratory results.<br>3. The performance of machine learning (ML) classifiers could be affected by the class proportions in the training dataset. The proportion of brain MRI reporting in AIS may vary significantly depending on the characteristics of each hospital. |
| 12 | NLP method successfully extracted the index lesion and its corresponding information from free-form text from breast MRI. | 1. The NLP system performed was not suitable for other reports that did not use BI-RADS descriptors.<br>2. Only 7 out of 478 cases reported no lesion in the data, and there is no case with BI-RADS 0 or 1 in the rest of |
|  |  | 471 reports which reflected the general population who underwent breast MRI.<br>3. The NLP system extracted the index lesion based on the authors' hypothesis that the index lesion accounts for the largest number of imaging features. This hypothesis was artificially defined and was not the direct extraction of the index lesion. |
| 13 | Radiology-arthroscopy correlation can be automated for knee meniscal tears using NLP algorithms, which shows promise for education and quality improvement. | 1. Some studies without a disagreement between the knee MRI and arthroscopy reports would not be screened out by the NLP approach, which would then still require manual review.<br>2. Data from a single institution.<br>3. In analyzing MRI and arthroscopy correlation, there is a delay between the two studies and patients may develop new meniscal abnormalities in that time interval.<br>4. Different types of meniscal tears may have different management implications<br>5. The focus of the study is on meniscal tears, but other abnormalities may be clinically relevant for radiology-arthroscopy correlation and would require further labeling of training data. |
| 14 | Rule-based NLP is efficient approach for identifying patients with Type 1 Modic change if the emphasis is on identifying only relevant cases with low concern regarding false negatives | 1. Difficulty of eliciting all possible keywords given the enormous variability of how lumbar spine findings are reported.<br>2. Limited number of reports for a specific finding, findings that are complex to identify, ambiguity in reports, and feature sets which are not sufficiently rich. |
|  |  | 3. F1 scores are measures that are limited to being a relative term with no absolute range or ranges of poor, fair, good, or excellent.<br>4. The developing rule-based algorithms is determining how far apart key words can be in a sentence to be considered together. |
| 15 | The developed model used labels extracted from radiology reports to provide automated diagnosis categorization of cardiac MR images with a high level of performance. | 1. The BERT model was not 100% accurate for all five diagnoses.<br>2. Researchers can use their model.<br>3. They did not explore alternative ways to overcome the 512-token input limitation of BERT. |
| 16 | Proposed NLP pipeline is capable of inferring BT-RADS report scores from unstructured reports after training on structured report data. The study provides a detailed experimentation process and may provide guidance for the development of RADS-focused information extraction (IE) applications from structured and unstructured radiology reports. | 1. It is a single-center, retrospective study.<br>2. The grouping of BT3 subcategories raises another limitation of the study, as the distinction between them is clinically important, differentiating between pseudo-progression and likely true tumor progression.<br>3. Some Model performed on unstructured reports. |
| 17 | Among various NLP techniques, the bag-of-words approach combined with a LASSO regression model demonstrated the best overall performance in extracting binary outcomes from free-text clinical reports. This study provides a framework for the development of machine learning-based | 1. A consensus in human classification was used as ground truth, which is a commonly used method to generate an approximation in the absence of actual ground truth.<br>2. Complete data set was manually classified to generate labels for training and testing, however when an NLP model will be used, only a minor portion will be labeled manually to predict the labels on the remaining data set.<br>3. Models trained on data from single institutions. |
|  | NLP models as well as a clinical vignette of patients diagnosed with brain metastases. |  |
| 18 | NLP demonstrated good accuracy for tumor status classification and may have novel application for automated disease status classification from electronic databases. | 1. limitations exist for unbalanced datasets where class sizes differ significantly.<br>2. As imaging features and terminology vary between different tumor types and imaging modalities, the algorithm may not be applicable to different patient populations.<br>3. The lack of uniformity across reports made complete alignment impossible. |
| 19 | The NLP system with rule-based pattern matching achieved rapid and accurate structured processing of rectal cancer MRI reports. MRI reports with structured templates are more suitable for NLP-based extraction of information. | 1. The dataset came from a single center.<br>2. MRI reports were based on the reading radiologists' interpretations, and the accuracy of the test dataset can be expected among the radiologists.<br>3. The low reporting rate of all image features in reports before 2015 may have resulted in a lack of representativeness. |
| 20 | Rule-based processing is an accurate method for the large-scale, automated extraction of PI-RADS scores from the text of radiology reports. These natural language processing approaches can be used for future initiatives in quality improvement in prostate mpMRI reporting with PI-RADS. | 1. RegEx algorithm was developed based on the reporting characteristics of a single institution.<br>2. RegEx algorithm was unable to categorize prostate mpMRI reports in 5.49% of cases.<br>3. The algorithm is unable to assess the underlying accuracy of the PI-RADS score assigned during the original clinical interpretation. |
| 21 | The NLP algorithm demonstrates high recall and precision for information | 1. A large amount of the breast MRI reports was selected from their department and thus the segmentation rules |
|  | extraction from free-text reports. This approach will help to narrow the gap between unstructured report text and structured data, which is needed in decision support and other applications. | and the reviewed BI-RADS lexicon were developed based on the writing habits of single department.<br>2. There are often multiple lesions in breast MRI reports and the index lesion is most crucial to clinicians in determining the management and prognosis of patients. However, this study extracted information from all the lesions in breast MRI reports not just from the index lesions. |
| 22 | A deep learning model based on CNN enables radiologists to discriminate between high – grade glioma (HGG) and metastasis based on MRI reports with high-precision values. | 1. insufficient number of radiology reports selected for the training and testing of the NLP tools .<br>2. the translation from Spanish to English reports would have some kind of impact on the outcome of language of NLP tool as linguistic nuances are probably being missed during the translation process.<br>3. lack of additional lesions on the radiology other than glioma. |
| 23 | The two NLP models developed accurately predict spine and head MRI protocol assignment, which could improve radiology workflow efficiency. | 1. NLP models determine protocol assignment by word and word-context relationships. which can lead to unintended use of non-medically relevant, human biases hidden in the data.<br>2. The head MRI protocol data lacked sufficient sample size on more specialized protocols.<br>3. Data comes from a single academic institution. |
| 24 | NLP models developed from the narrative clinical information provided by referring clinicians and demographic data are feasible | 1. Data comes from a single academic institution.<br>2. For most of the study period, requisitions were completed by hand and the study relies on the faithful reproduction |
|  | methods to predict the protocol and priority of MRI brain examinations. | of handwritten text by administrative staff or the interpreting radiologist<br>3. Absence of important inputs like patient allergy information, glomerular filtration rate, or diabetes status, must rely on the referring service and age of the patient to predict the administration of gadolinium.<br>4. Although protocol guidelines exist at their institution, it is not possible to completely remove variation in protocol selection.<br>5. Due to the limitations of the picture archiving and communication system, a smaller subset of the dataset was used to evaluate the priority model. |
| 25 | A natural language classification algorithm can be trained with IBM Watson to automatically determine the need for intravenous contrast in musculoskeletal MRIs. | 1. Challenges with spelling, grammar, and ambiguity in clinical indications.<br>2. Difficulty troubleshooting errors due to a "black-box" algorithm.<br>3. Issues with contrast assignment, potentially leading to clinical consequences.<br>4. Lack of consideration for the requested study type, such as "MRI lumbar spine without contrast."<br>5. Constraints in accessing and modifying IBM Watson's closed cloud service.<br>6. Risks of harmful errors from potential updates to the service's algorithm.<br>7. An intrinsic limitation in the scalability of our methods at the institution was the assignment of MRI protocols (which serves as ground truth) as free-text. |
| 26 | Application of NLP-based techniques has the potential to significantly reduce the time and cost spent on protocoling appropriate examinations. The results indicate that the proposed model can automate the assignment of orders for further revision. The low number of false negative instances suggests the reliability of the model | 1. Limited number of protocols were implemented.<br>2. Limitation in the generalizability of the model.<br>3. The cost analysis is solely based on economic feasibility that is not considering the costs associated with potential patient harm and delay in treatment due to a false negative. |
| 27 | NLP-based deep learning (DL) algorithms can be used as an important digital marker for unstructured electronic health record data DL prediction. | 1. MRI text report was read by neuroradiologists in single institution.<br>2. They couldn't conclude whether these DL algorithms will perform better in predicting poor outcomes using brain MRI text reports in languages other than English. |
| 28 | The deep-transfer-learning model using free-text radiological reports can predict the survival of patients with rectal cancer, thereby increasing the utility of unstructured medical big data. | It was a retrospective study conducted at a single institution. |
| 29 | AI-based data extraction and analysis tools are becoming more available and bringing new insights into unstructured health data records. This study shows that this approach is feasible and reveals plausible data. With this they were able to confirm the predictive value of SP-CMR for CAD assessment. Positive ischaemic test and the presence of late gadolinium enhancement ( LGE) was | 1. A significant number of cases were excluded thus it would be expected to inflate the importance of CMR measures and produces some bias of the results.<br>2. The NLP model in this study was not exclusive and some data extraction was performed using CogStack search engine of instances in texts and structured fields. |
|  | associated with a higher risk of mortality regardless of clinical risk factors. | CMR reports at the time of this study were not included in NLP training and manual extraction was performed.<br>Single large center study and only tested on one population. |
| 30 | bidirectional encoder representations from transformers, graph convolutional network [BERT][GCN] was externally validated to predict isocitrate dehydrogenase (IDH) mutation status in patients with diffuse glioma using routine MR radiology reports with superior or at least comparable performance to human reader. | There was a difference in the proportion of patients with IDH mutation between the two hospitals used for model development and patients with IDH mutation in the external validation higher proportion of set. This distribution shift may hamper fair performance evaluation of the models.<br>The radiology reports since the introduction of electronic health record at the respective hospitals were included dating back to 2009 when the relevance and prognostic implication of IDH mutation were just being reported IDH mutation was incorporated in WHO classification of central nervous system tumor in 2016 with increasing efforts to associate imaging findings to molecular markers.<br>sample size calculation was not performed when designing this study.<br>While long short-term memory (LSTM) enables estimation of relative importance of text variables, it still remains largely unknown the based models extract and synthesize -exact mechanism transformer information on free texts. |
| 31 | NLP can identify patterns of lumbar spine degeneration through analysis of a large corpus of radiologist interpretations. Demographic differences in stenosis | The NLP algorithm empowers a much larger corpus to be analyzed, but its classification is imperfect. |
|  | prevalence shed light on the natural history and pathogenesis of Lumbar spine degenerative disease( LSDD). | 2. Their rules-based algorithm relied on a manually assembled dictionary mapping non-standard terminology for stenosis grading to A 6-point standard scale. This methodology is imperfect and may affect our estimate of the severity distribution of LSDD.<br>3. The study cohort is not a true sampling of the general population as only patients with symptoms substantial enough to warrant imaging are studied. |
| 32 | GPT-4 demonstrates the potential to provide largely accurate and clinically useful treatment recommendations for common orthopedic knee and shoulder conditions. Expert surgeons rated the recommendations at least as "good", but the patient's situation and treatment urgency were not fully considered. Therefore, patients need to consult healthcare professionals for personalized treatment recommendations, while GPT -4 may be a supplementary resource rather than a replacement for professional medical advice after regulatory approval. | 1. They studied only a few patients.<br>2. To enhance its depth and relevance to clinical scenarios, GPT-4's predictions need to be more specific.<br>3. The patient spectrum was broad.<br>4. Treatment recommendations were qualitatively judged by two experienced orthopedic surgeons, and involvement of more surgeons could have strengthened the outcome basis even further.<br>5. The tendency of GPT-4 to give generic and unspecific answers and to err on the side of caution rendered it challenging to assess its adherence to guidelines or best practices exactly.<br>6. They used a standardized and straightforward way of prompting GPT-4. After more extensive modifications of these prompts, outcomes may be different. |
| 33 | Artificial intelligence approaches such as expert-driven NLP and ontology-driven NLP have similar accuracy in identifying | 1. Unavailability of actual clinical outcomes for radiology department with evaluation.<br>2. The study was limited to clinical notes obtained from single academic medical center. |
|  | patients with implantable devices that pose high safety risks for MRI. |  |
| 34 | A logistic regression model was created to predict which patients may require spine surgery. Simple clinical variables appeared more predictive than variables created using NLP. | 1. A low sample size for this type of analysis.<br>2. Data came from one spine assessment clinic, and the model has not been externally validated.<br>3. By design, it relies on retrospective data, which can be prone to misclassification bias.<br>4. The surgical outcomes of the patients in the study are unknown, thus it's not clear whether the surgical decisions made were optimal. |
| 35 | Application of ChatGPT to summarize MRI reports at a reading level appropriate for patients. Physicians were likely to be satisfied with the summarized reports with respect to factual correctness, ease of understanding, and completeness. Physicians were less likely to be satisfied with respect to potential for harm, overall quality, and likelihood to send to patients. | 1. Small sample size and a single institution study.<br>2. They focused exclusively on prostate cancer MRI reports, and the results may not be directly applicable to other types of radiology reports or cancer diagnoses.<br>3. The anonymous questionnaire used in the study may be subject to response bias, as physicians with strong opinions on AI-generated summaries may be more likely to participate.<br>4. There were concerns regarding "potential for harm, overall quality, and likelihood of sending the report to patients. The study is unable to identify potential causes for this discrepancy, and it is beyond the current scope of the study.<br>5. It's possible that physicians communicated results with patients and did not document this in the electronic medical record. |

NLP was found to have diverse applications in research studies, predominantly in staging, quantification, disease diagnosis, and protocol selection.

Seven studies have focused on employing *NLP for staging and quantification*. Lee SJ et al. ^16^ demonstrated the accurate interpretation of BT-RADS report scores through NLP trained on structured reports, while Zhang D et al. ^20^ utilized a rule-based algorithm to automatically categorize prostate MRI reports.

Six studies utilized NLP for *disease diagnosis*. Notably, in the studies by Dewaswala N et al. ^10^ and Liu Y et al.^12^, NLP effectively extracted diagnoses like hypertrophic cardiomyopathy and information about index lesions from breast MRI reports, respectively.

Moreover, NLP was integrated into four studies for *protocol selection*, exemplified by Trivedi H et al. ^25^ study, which employed natural language classification to identify contrast requirements in musculoskeletal MRIs.

In the realm of *prognosis prediction*, Kim S et al.^28^ introduced a novel computer model to analyze MRI reports of rectal cancer patients for estimating their survival times.

Additionally, Valtchiney VI et al. ^33^ explored methods for *identifying MRI safety risks* related to implanted devices in a study focusing on safety protocol compliance, while Chung EM et al.^35^ not only developed a logistic regression model to predict patients requiring spine surgery but also investigated the use of ChatGPT technology to succinctly *summarize MRI reports* and enhance patient comprehension.

## Discussion

Our review showcased the growing importance of NLP in MRI, with 26 publications covering over 160k MRI reports, across several organ systems. However, a key weakness is that only one prospective study was conducted ^27^, such studies are essential for validating NLP algorithms in real-time clinical and operational settings to bolster clinical decision-making, workflow efficiency, and personalized medicine approaches. Future research should focus more on prospective studies to better validate the real-world benefits.

Additionally, recommendations include expanding NLP applications to other radiology fields such as pediatric radiology, and particularly in the context of cancer diagnosis and staging to broaden the impact and potential utility of NLP in the field.

The results of our review highlight several significant opportunities to streamline MRI imaging processes using advanced technology. By implementing more sophisticated protocols, bolstered by NLP, we could potentially reduce the time required for approving referrals ^26^. This simplification means a more efficient workflow for healthcare providers.

Additionally, our findings suggest that NLP can simplify interpretative data for patients. This transparency allows patients to better understand their health information, which can improve their engagement and satisfaction with the treatment process ^35^.

For radiologists, the technology we studied offers support in summarizing complex imaging results, such as those from MRI scans. This aid not only speeds up their workflow but also enhances the accuracy and comprehensiveness of the diagnostic data provided, especially in MRI reports with structured templates^19^.

Looking ahead, the possibilities for further integration of this technology across different imaging domains are vast. Each specialty can learn from the others, leveraging technological advancements for the benefit of patient care and system efficiency.

Our review has limitations. First, we did not perform a meta-analysis due to the high heterogeneity in the methodologies and tasks used across the referenced studies, which made direct comparisons problematic. Second, our review was limited to articles sourced from PubMed, potentially omitting relevant studies published in other databases. Furthermore, the scope of our analysis was restricted to English-language publications, excluding potentially significant research available in other languages. Lastly, our findings are confined to the data available up to the point of our review, and as such, newer studies post-review are not considered.

In conclusion, NLP applications in MRI show potential for a change in the field. However, while the review revealed a wealth of evidence supporting the effectiveness of NLP in MRI analyses, the presence of just one prospective studies underscores the need for further research to validate NLP algorithms in real-time clinical and operational settings.

Moreover, expanding the scope of NLP usage to encompass other radiology specialties, presents opportunities for advancing healthcare practices. By addressing these recommendations in future research endeavors, the integration of NLP technologies stands to enhance clinical decision support, drive research advancements, and improve patient outcomes in the realm of MRI imaging.

## Data Availability

All data produced in the present study are available upon reasonable request to the authors

