## Supplemental 1 for "Systematic review of natural language processing (NLP) applications in magnetic resonance imaging (MRI)"

### Supplementary Online Content

***Supplementary Material 1.*** Literature search strategy.

***Supplementary Table 1***. Quality Assessment of Diagnostic Accuracy Studies-2.

#### ***Supplementary Material 1: literature search strategy***

A comprehensive literature search was performed to identify studies evaluating the clinical applications of NLP for MRI. The search was conducted on January 4, 2024 , using the PubMed database.

Search keywords included " MRI ", " Magnetic resonance imaging", MRE", " Magnetic resonance enterography", " NLP", "Natural Language Processing ", "LLM", "large language models", and "chatGPT" . Details on complete search strategies are provided in Supplementary Material.

**Supplementary Table 1: Quality Assessment of Diagnostic Accuracy Studies-2 (QUADS-2) risk of bias assessment.**

Abbreviations: Pt. patient; Ref. reference. ✗ = high risk of bias; ✓ = low risk of bias.

[a]Failing to describe their study population; [b]external validation; [c]Who performed the annotations? [d]Did all patients receive the same reference standard? Were all patients included in the analysis? [e]Failing to specify ethical approval.

| Author | Risk of bias | | | | |
|---|---|---|---|---|---|
| | Pt. selection[a] | Index test[b] | Ref. standard[c] | Flow and timing[d] | Data management[e] |
| Dewaswala N, Arruda-Olson AM.[1] | ✓ | ✓ | ✓ | ✓ | ✓ |
| Kim C, Lenert L.[2] | ✓ | ✓ | ✗ | ✓ | ✓ |
| Kim M, Sohn B.[3] | ✓ | ✓ | ✓ | ✓ | ✓ |
| Lee SJ, Banerjee I.[4] | ✓ | ✓ | ✓ | ✓ | ✓ |
| Senders JT, Arnaout O.[5] | ✓ | ✓ | ✓ | ✓ | ✓ |
| Liu Y, Wang X. [6] | ✓ | ✓ | ✓ | ✓ | ✓ |
| Cheng LT, Erickson BJ.[7] | ✓ | ✓ | ✓ | ✓ | ✓ |
| Li MD, Huang AJ. [8] | ✓ | ✓ | ✓ | ✓ | ✓ |
| Chillakuru YR, Sohn JH.[9] | ✓ | ✓ | ✓ | ✓ | ✓ |
| Liu W, Li Y. [10] | ✓ | ✓ | ✓ | ✓ | ✓ |
| Huhdanpaa HT, Jarvik JG.[11] | ✓ | ✓ | ✓ | ✓ | ✓ |
| Brown AD, Marotta TR.[12] | ✓ | ✓ | ✓ | ✓ | ✓ |
| Zhang D, Gupta RT.[13] | ✓ | ✓ | ✓ | ✓ | ✓ |
| Travis Caton M Jr, Andriole KP.[14] | ✓ | ✓ | ✗ | ✓ | ✓ |
| Liu Y, Wang XY.[15] | ✓ | ✓ | ✓ | ✓ | ✓ |
| Trivedi H, Sohn JH.[16] | ✓ | ✓ | ✓ | ✓ | ✓ |
| Truhn D, Nebelung S.[17] | ✓ | ✓ | ✓ | ✓ | ✓ |
| Valtchinov VI, Khorasani R.[18] | ✓ | ✓ | ✓ | ✓ | ✓ |
| Heo TS, Kim C.[19] | ✓ | ✓ | ✓ | ✓ | ✓ |
| Kim S, Shin SJ.[20] | ✓ | ✓ | ✓ | ✓ | ✓ |
| Eghbali N, Ghassemi MM.[21] | ✓ | ✓ | ✓ | ✓ | ✓ |
| Alskaf E, Chiribiri A.[22] | ✓ | ✓ | ✓ | ✓ | ✓ |
| Martín-Noguerol T, Luna A.[23] | ✓ | ✓ | ✓ | ✓ | ✓ |
| Krebs B, Gross DP.[24] | ✓ | ✗ | ✓ | ✓ | ✓ |
| Zaman S, Linton N.[25] | ✓ | ✓ | ✓ | ✓ | ✓ |
| Chung EM, Kamrava M. [26] | ✓ | ✓ | ✓ | ✓ | ✓ |

#### Refrences
